# The Dissemination of Multidrug-Resistant and Hypervirulent *Klebsiella pneumoniae* Clones Across the Kingdom of Saudi Arabia

**DOI:** 10.1101/2024.03.26.24304793

**Authors:** Jiayi Huang, Ahmed Yousef Alhejaili, Usamah Hussein Alkherd, Mathew Milner, Ge Zhou, Deema Alzahrani, Manuel Banzhaf, Albandari A. Alzaidi, Ahmad A. Rajeh, Maram Abdulmohsen Al-Otaiby, Sara Binabbad, Doua Bukhari, Abdullah N. Aljurayan, Alanoud T. Aljasham, Zeyad A. Alzeyadi, Sulaiman M. Alajel, Pei-Ying Hong, Majed Alghoribi, Mashal M. Almutairi, Arnab Pain, Waleed Al Salam, Danesh Moradigaravand

**Author notes:** Shared first/last authorship (Jiayi Huang, Ahmed Yousef Alhejaili, Waleed Al Salam, Danesh Moradigaravand).

## Abstract

The Gram-negative bacterium *Klebsiella pneumoniae* is a major human health threat underlying a broad range of community- and hospital-associated infections. The emergence of clonal hypervirulent strains often resistant to last-resort antimicrobial agents has become a global burden as treatment options are limited. The Kingdom of Saudi Arabia (KSA) has a dynamic and diverse population and serves as a major global tourist hub facilitating the dissemination of multidrug-resistant (MDR) strains of *K. pneumoniae*. To examine the spread of clinically relevant *Klebsiella pneumoniae* strains, we characterized the population structure and dynamics of multidrug-resistant *K. pneumoniae* across the KSA hospitals.

**Methods:** We conducted a large genomic survey on a Saudi Arabian collection of multidrug-resistant *K. pneumoniae* isolates from bloodstream and urinary tract infections. The isolates were collected from 32 hospitals located in 15 major cities across the country in 2022 and 2023. We subjected 352 *K. pneumoniae* isolates to whole-genome sequencing and employed a broad range of genomic epidemiological and phylodynamic methods to analyse population structure and dynamics at high resolution. We employed an integrated short- and long-read sequencing approach to fully characterize multiple plasmids carrying resistance and virulence genes.

**Results:** Our results indicate that, despite a diverse *K. pneumoniae* population underlying hospital infections, the population is characterized by the rapid expansion of a few dominant clones, including ST096 (n=115), ST147 (n=75), ST231 (n=35), ST101 (n=30), ST11 (n=18), ST16 (n=15) and ST14 (n=12). ST2096, ST231, and ST147 clones were estimated to have formed within the past two decades and spread between hospitals across the country on an epidemiological time scale. All STs were genetically linked with globally circulating clones, particularly strains from the Middle East and South Asia. All of the major clones harboured plasmid-borne ESBLs and a range of carbapenemase genes. Plasmidome analysis revealed multiple mosaic plasmids with resistance and virulence gene cassettes, some of which were shared between the major clades and account for multidrug resistant hypervirulent (MDR/hv) phenotype, especially in the ST2096 strains. Integration of phylodynamic data and resistance plasmid profiles showed that the acquisition of plasmids occurred on the same time scales as did the expansion of major clones across the country.

**Conclusion:** Taken together, these results indicate the dissemination of MDR and MDR-hv *K. pneumoniae* strains across the kingdom and provide evidence for pervasive plasmid sharing and horizontal gene transfer of resistance genes. The results demonstrate the independent introduction of endemic ST147, ST231, and ST101 clones into the country and highlight the clinical significance of ST2096 as an emerging clone with dual resistance and virulence risks. These results highlight the need for continuous surveillance of circulating and newly emergent strains (STs) and of their plasmidome footprints carrying MDR determinants.

## Introduction

*K. pneumoniae* is a genetically diverse Gram-negative bacterium implicated in several hospital-acquired infections, including urinary tract and bloodstream infections [1]. Pathogenic strains of *K. pneumoniae* colonize a broad range of niches. These strains have a dynamic genome in which key clinically significant genes for resistance and virulence can be acquired, leading to the emergence of high-risk strains from nonpathogenic strains [2]. The emergence and global spread of strains resistant to last-resort carbapenem antimicrobials over recent decades have been recognized as a health priority by the World Health Organization, which places *K. pneumoniae* on the priority list of pathogens for which imminent therapeutic solutions are required [3].

Elevated virulence and resistance levels of *K. pneumoniae* are associated with mortality and poor clinical outcomes. Clones of *K. pneumoniae* emerge locally within hospitals and spread regionally and globally [4]. These strains are commonly recovered from various distant geographical locations, can colonize, or infect multiple hosts and are rapidly transmitted across hosts and settings. The best-characterized international clones are CG258, which include ST258, S11, clonal groups CG14/15, CG17/20, CG43, CG147, and ST101 [4–6]. In recent decades, these clades have converged between carbapenem resistance and virulence [7]. Multidrug resistant (MDR) and hypervirulent (hv) strains, in which nonoverlapping genomic signatures for resistance cooccur in the same genomic context, are prevalent and are a cause for concern in clinical settings [7, 8]. These strains can cause severe infections within hospital settings but also within healthy individuals in the community, with very limited treatment options [9]. The strains showed a broad phylogenetic distribution and were rapidly disseminated across countries and healthcare systems.

The Kingdom of Saudi Arabia (KSA) is strategically located and has an expansive geographical area and a complex and dynamic demography. The country is also an international hub for travel, trade, and religious tourism, hosting mass gathering events. These factors have turned the country into a potential melting pot for the import and transmission of antimicrobial-resistant strains worldwide. This setting also allows for the horizontal transfer of resistance and virulence genes. High carbapenem usage in the kingdom is associated with a rapid increase in the incidence of carbapenem-resistant Enterobacterales (including *K. pneumoniae*). Multiple small-scale studies on individual or small networks of hospitals have reported the incidence of MDR clones in the country [10–14]. In these studies, classical molecular typing methods with poor resolution or heterogeneous and inconsistent datasets from different time points have mainly been employed. Therefore, the dynamics of resistant clones and their resistance mechanisms remain largely understudied.

To address this gap, we conducted a large-scale precision epidemiology analysis to decipher the epidemiology and evolution of multidrug-resistant *K. pneumoniae* strains in the country. We curated a collection of samples systematically recovered from 34 hospitals across the country from bloodstream and urinary tract infections. The depth and breadth of the sampling provided us with the most comprehensive picture of the clones circulating in the country during a period of one and a half years. We fully characterized the diverse population of *K. pneumoniae*, including the prevalent clones in the country and their plasmid content, which are responsible for resistance, virulence and convergence of resistance and virulence. Our study revealed the dissemination of both globally and regionally endemic clones and the extensive sharing of plasmids associated with virulence and resistance between the major circulating clones.

## Methods

### Sampling and collection

This study was approved by Institutional Review Board (IRB) of King Abdullah University for Science and Technology (approval number 23IBEC027) and IRB of Saudi Ministry of Health (approval number: 23-23 M). As part of the hospital surveillance program, 352 *K. pneumoniae* isolates and their epidemiological data were collected between January 2022 and April 2023. Samples were retrieved from the bloodstream and urinary tract infections. For samples from bloodstream infections, we obtained clinical data for death and cause of death. We isolated strains from patients who were infected with MDR *K. pneumoniae*, that is, who were resistant to more than two classes of antimicrobials, via analysis in a molecular microbiology laboratory in the hospital. A combination of phenotypic and genotypic methods was used to identify the MDR *K. pneumoniae* strains. This included antimicrobial susceptibility testing (AST) and polymerase chain reaction (PCR) to identify the *K. pneumoniae* and β-lactamase genes. To explore the links with nonclinical reservoirs, we included nine strains recovered from retail food (lettuce) in Jeddah [15]. We also integrated our collection with the genomic collection from a single hospital study in Jeddah for phylodynamic analysis [16].

### Strain identification and susceptibility testing

The Vitek-2 compact was used for identification and antibiotic susceptibility testing (BioMerieux, Marcy-l’Etoile, France). ID-GN (reference: 22226) and GN-AST-291 (reference: 415062) cards were used for identification and antibiotic susceptibility testing, respectively. Briefly, the isolates were cultured overnight on agar plates at 37 °C. A 0.45% sodium chloride (NaCl) solution (CareFusion, USA) was used to prepare a bacterial suspension with an optimal density of 0.5–0.63 McFarland units. The VITEK cards were inoculated and loaded in the Vitek-2 compact according to the manufacturer’s instructions. The system output was generated automatically. The GN-AST-291 card contains 35 antibiotics: ESBL, amoxicillin (AMX), ampicillin (AMP), amoxicillin/clavulanic acid (AMC), piperacillin/tazobactam (PTZ), cefalotin (CEF), cefazolin (CFZ), cefuroxime (CXM), cefoxitin (FOX), cefixime (CFM), ceftazidime (CAZ), ceftizoxime (CTZ), ceftriaxone (CRO), cefepime (FEP), ertapenem (ETP), imipenem (IMP), meropenem (MEM), amikacin (AMK), gentamicin (GEN), tobramycin (TOB), ciprofloxacin (CIP), tigecycline (TGC), nitrofurantoin (NIT), trimethoprim (TMP), cefotaxime (CTX), and piperacillin/sulbactam (TZP).

### Short-read sequencing

*K. pneumoniae* strains were cultured overnight at 37°C in Luria–Bertani (LB) broth. Genomic DNA (gDNA) was extracted using a DNeasy Blood and Tissue Kit according to the manufacturer’s instructions (QIAGEN, Hilden, Germany). The quality of the gDNA was determined using a DS-11 DNA quantification spectrophotometer from Denovix (Denovix, US). The gDNA quantity was determined using a fluorometric method with analysis performed on a Qubit 4.0 fluorometer; a high-sensitivity double-stranded DNA assay kit (Thermo Fisher Scientific, US) was employed. We then prepared genomic libraries for the 361 isolates using the MGIEasy Fast FS DNA Library Prep in accordance with the instructions in the user manual provided by MGI Technology (MGI Technology, China). Therefore, an enzymatic fragmentation protocol was used. The libraries were then denatured and circularized. Whole genome sequencing (WGS) was performed on the DNBSEQ-G400 platform (MGI Technology, China) using the 2 x 150 bp paired-end read protocol.

The short reads were subjected to quality control with the fastqc package in R (v0.1.3). We assembled the genomes with a *de novo* assembly pipeline using Unicycler (v0.5.0) (https://github.com/rrwick/Unicycler#install-from-source) with default parameters [17]. Assemblies were processed, and contigs shorter than 200 bp were excluded from the analysis with Unicycler. We profiled and characterized the strains with Kleborate (v2.3.2) to determine the sequence and capsule types and the resistome and virolome of the collection [18]. The risk assessment scores for resistance and virulence levels were also obtained for Kleborate. Virulence scores, graded on a scale of 0 to 5, are computed by the detection of specific genetic markers linked to heightened pathogenicity, with yersiniabactin (*ybt*) having the smallest risk, followed by colibactin (clb), and then aerobactin (*iuc*). The presence of the loci *ybt, ybt+clb, iuc, ybt+iuc and ybt+clb+iuc* corresponded to virulence scores of 1 to 5. The absence of all of these loci corresponded to virulence score of zero. Similarly, resistance scores, ranging from 0 to 3, were determined by the presence of antimicrobial resistance genes, with ESBL being the least resistant, followed by carbapenemase, and then carbapenemase combined with colistin resistance determinants. The convergence of resistance and virulence occurs when both virulence and resistance scores were equal to or greater than 3 and 1, respectively [18].

Virulence markers and resistance determinants were identified using srst2 v0.2.0 (accessible at www.github.com/katholt/srst2) [19] against the PlasmidFinder (v2.1.1) [20], VFDB (v6.0) [21], ResFinder (v4.1) [22], and CARD (v3.2.8) [23] databases. A threshold of 90% identity was applied to detect genes. We annotated *de novo* assemblies with Prokka (v1.14.5) [8] and fed the annotated assemblies into Panaroo to reconstruct the pangenome [9]. To generate the phylogenetic tree for our dataset, we initially aligned the short-read sequences against the genome of the reference strain *K. pneumoniae* Ecl8, an ST375/K2 hypervirulent clone of significant epidemiological importance in causing severe community-acquired infections (accession number: PRJEB401) [24]. This alignment was conducted using the Snippy pipeline (accessible at https://github.com/tseemann/snippy) with default settings. We then calculated the pairwise SNP distances among the core genome alignments and used the FastTree package (2.1) [25] with default parameters to construct an approximately maximum likelihood phylogenetic tree.

### Transmission analysis and phylodynamic analysis

We combined our genomes with those sequenced in a single hospital study in Jeddah [16], to attain a larger collection size. For the five prevalent sequence types (STs), i.e., STs with more than 1% of the population—ST14, ST101, ST231, ST147, and ST2096—within the two collections combined, we conducted phylodynamic analysis to unveil the crucial epidemiological characteristics of each clone and reconstruct dated phylodynamic trees. We exclusively considered strains within each ST group with collection dates for our analysis.

We selected the strain with the best assembly statistics, namely, the highest N50, for each clone. We merged the contigs of these selected strains to form a local reference genome. By aligning the short reads of each strain in the clone to this reference genome, we derived a core genome SNP alignment. This alignment was fed into Gubbins (v.3.3.1) [12] with five iterations to eliminate hypervariable regions. The SNP counts for the major clones were as follows: ST101 (1,125 sites), ST231 (579 sites), ST147 (1,837 sites), ST2096 (2624 sites), and ST14 (1,923 sites). To estimate transmission links, we assumed a SNP difference cut-off of less than 10 for a transmission event. Given the mutation rate of 5 SNPs/year for *K. pneumoniae*, this cut-off corresponded to events over two years.

To estimate the ancestral locations of the major clones within the SNP clusters, we performed phylogeographic diffusion analysis in discrete space using BEAST (v2) [26]. We used the geographical location (city of isolation) as a discrete state of each taxon and employed a constant population size model with uniform priors on the clock rate. We also utilized a symmetric model with a uniform prior distribution for the discrete trait substitution model for the diffusion of the clone in space. Convergence of the Markov chain Monte Carlo (MCMC) chains was assessed by ensuring that the effective sample size (ESS) for key parameters exceeded 150. The spatial trajectories derived from the dated Bayesian tree were visualized using the www.spreadviz.org website.

For a thorough examination of population size changes over time, we employed a nonparametric growth model to sample population sizes on the dated phylogenetic tree for the ST2096, ST231 and ST147 clones. We therefore used the skygrowth.mcmc function and visualized the results with the plot function within the skygrowth package (v0.3.1) [27].

### Contextualization of the samples

We contextualized our strains using the Pathogen Detection database available at https://www.ncbi.nlm.nih.gov/pathogens/. We first retrieved epidemiological SNP clusters including the strains from the Pathogen Detection database on 06/11/2023. These SNP clusters contained all strains with pairwise SNP distances of up to 50 SNPs. The clustering process was conducted by the pipeline within the Pathogen Detection portal (https://www.ncbi.nlm.nih.gov/pathogens/pathogens_help/#references). This high-throughput automated pipeline employs a combined kmer approach and alignment to define clusters, with a fixed cut-off of 50 SNPs that cannot be customized. Subsequently, we extracted the trees for selected clusters from the portal and visualized them with the ggtree package (v3.8.2) in R [28].

### Plasmidome analysis and long-read sequencing

We conducted third-generation sequencing on 20 representative samples from the population to fully resolve the plasmid content. As the plasmid content of the ST2096 strains was characterized before [16], we selected samples from the other STs. Samples were selected from the dominant clones with the highest resistance and virulence scores from different clades. To prepare sequenced libraries for long-read sequencing, we employed 96-plex Rapid Barcoding Kits for multiplexing. These libraries were then loaded into MinION flow cells (Oxford Nanopore Technologies) and subjected to a 48-hour run following the manufacturer’s protocol. Hybrid assembly was conducted using Unicycler, employing the conservative option. Subsequently, the assembled contigs were screened for the presence of full copies of the origin of replications, virulence factor genes, and antimicrobial resistance genes using BLAST in conjunction with the abovementioned databases. The assembled genomes were visualized and checked with Bandage (v0.9.0) [29]. We extracted plasmid fragments containing virulence factors and resistance genes. Plasmids were visualized and annotated with the built-in tools in the Proksee portal (www.proksee.ca) [30]. We identified resistance genes and plasmid replicons via BLAST searches of the extracted plasmid fragments on the CARD [23] and PlasmidFinder [20] databases, as described above. We mapped the short reads of the other strains against these extracted plasmid fragments and used the mapping coverage to confirm the presence of these plasmids in the rest of the population.

### Integration of resistome and virulome data with phenotypic and clinical metadata

We integrated the genetic profile of resistance determinants obtained from Kleborate with the phenotypic resistance data using the odds ratio function from the Epitools package (v0.5.10.1) in R. Moreover, we examined the significance of associations between the accessory genes and SNPs and the resistance phenotype while considering population structure by employing Scoary (1.6.16) [31] on the Paneroo output and the SNPs identified via postread mapping to the reference genome, respectively. Specifically, we focused on pairwise p values (both worst and best p values) that adjusted for the confounding effect of population structure (lineage effect). The associations were assessed using Scoary (accessible at www.github.com/AdmiralenOla/Scoary). To visualize the tree and associated metadata, we utilized the ggtree package (v3.8.2) in R [28].

We obtained clinical metadata on the reported mortality, cause of death, and date of in-hospital death for all patients with bloodstream infections and those with urinary tract infections, which subsequently led to bacteraemia. We computed the mortality rates at days 7, 14, 21 and 30, considering the period between sample isolation and reported death, for patients infected by each major ST. We also computed the odds ratio of death for infection by the major STs and strains with different profiles of resistance determinants using the Epitools package.

### Statistical significance tests

We used a one-way proportion test in R to assess the significance of the differences in the ratios. The one-way Wilcoxon signed-rank test was used to assess the significance of differences between the means. To compare the significance of different parameters inferred from the Bayesian analysis, we considered the 95% lower and upper bounds of the credible intervals (highest posterior density (HPD)), which is the shortest interval that includes 95% of the probability density.

### Data availability

The genomic data have been deposited in the European Nucleotide Archive (ENA) under the study accession number PRJEB66182. Moreover, the assemblies have been uploaded to the NCBI GenBank database under the accession number PRJNA1018815. Detailed metadata associated with the genomes are provided in Supplemental Table S1. Sample IDs in the table were not known to anyone outside the research group.

## Results

### The rate of *K. pneumoniae* infections and resistance level in the KSA

We analysed *K. pneumoniae* samples collected from infections across 34 hospitals through routine procedures in the Ministry of Health hospital networks of the KSA. The infection rate of *K. pneumoniae* was 33.51% of all Gram-negative ESKAPE infections during the period of the study, establishing the organism as a predominant pathogenic strain within the country. The rates of resistance to carbapenems, cephalosporins, ciprofloxacin, and tigecycline in *K. pneumoniae* infections were 32.60%, 51%, 9.01%, and 9.02%, respectively. Of the strains, we sequenced 200 samples from bloodstream infections and 152 from urinary tract infections that were identified as multidrug resistant (resistant to more than two classes of antimicrobials [32]). We also included nine samples recovered from food sources (lettuce).

### Globally and locally circulating sequence types (STs) underlie the *K. pneumoniae* population diversity

The studied population of *K. pneumoniae* in KSA hospitals was diverse and consisted of 35 different STs. Five STs, i.e., ST2096 (n=115), ST147 (n=75), ST14 (n=12), ST101 (n=30) and ST231 (n=35), ST307 (n=11), ST16 (n=15) and ST11 (n=18), individually represented more than 1% of the population, and in total, they comprised 88% of the population (Figure 1A, Supplementary Table S1). The STs reported in a single-hospital study in Jeddah [16], were shared with the strains under study from Saudi cities, showing that the population diversity of strains in the hospital reflects countrywide diversity. ST2096, ST14, ST101, ST147 and ST231were found the most frequent types across the two collections combined and therefore we focus on these STs. The K-locus diversity showed that ST2096, ST14, ST101 and ST231 contained predominantly one type, i.e., KL64, KL2, KL17 and KL51, respectively, while for ST147, three KL2 (n=50), KL51 (n=3), and KL10 (n=9) types were observed, suggesting the copresence of multiple sublineages of ST147 in hospitals. Although ST11 and ST258 are commonly reported as the dominant multidrug-resistant hypervirulent *K. pneumoniae* strains globally, only 5% of the strains in our collection were ST11, and no ST258 cases were observed in our study cohort. The results indicate a somewhat distinctive pattern of *K. pneumoniae* epidemiology in the KSA, in which unlike the global population non-GS258 (K1/K2) strains are predominant. ST147 has a widespread distribution in multiple geographical areas, while ST231 is prevalent in South and East Asia [8]. ST2096 has not been widely reported as a globally circulating clone and mainly reported from the Middle East and South Asia [8, 33]. The phylogenetic tree of *K. pneumoniae* in the KSA also revealed a diverse population composed of distinct lineages and expanding STs (Figure 2). The largest clone population belonged to the CC14 strains, i.e., the ST14 and ST2096 clades, similar to recent reports from hospitals in the Middle East [34, 35] and from a single hospital analysis in Jeddah [16]. Moreover, clinical strains from various sources, i.e., body sites, and regions show an even phylogenetic distribution across lineages (Figure 2).

**Figure 1.**
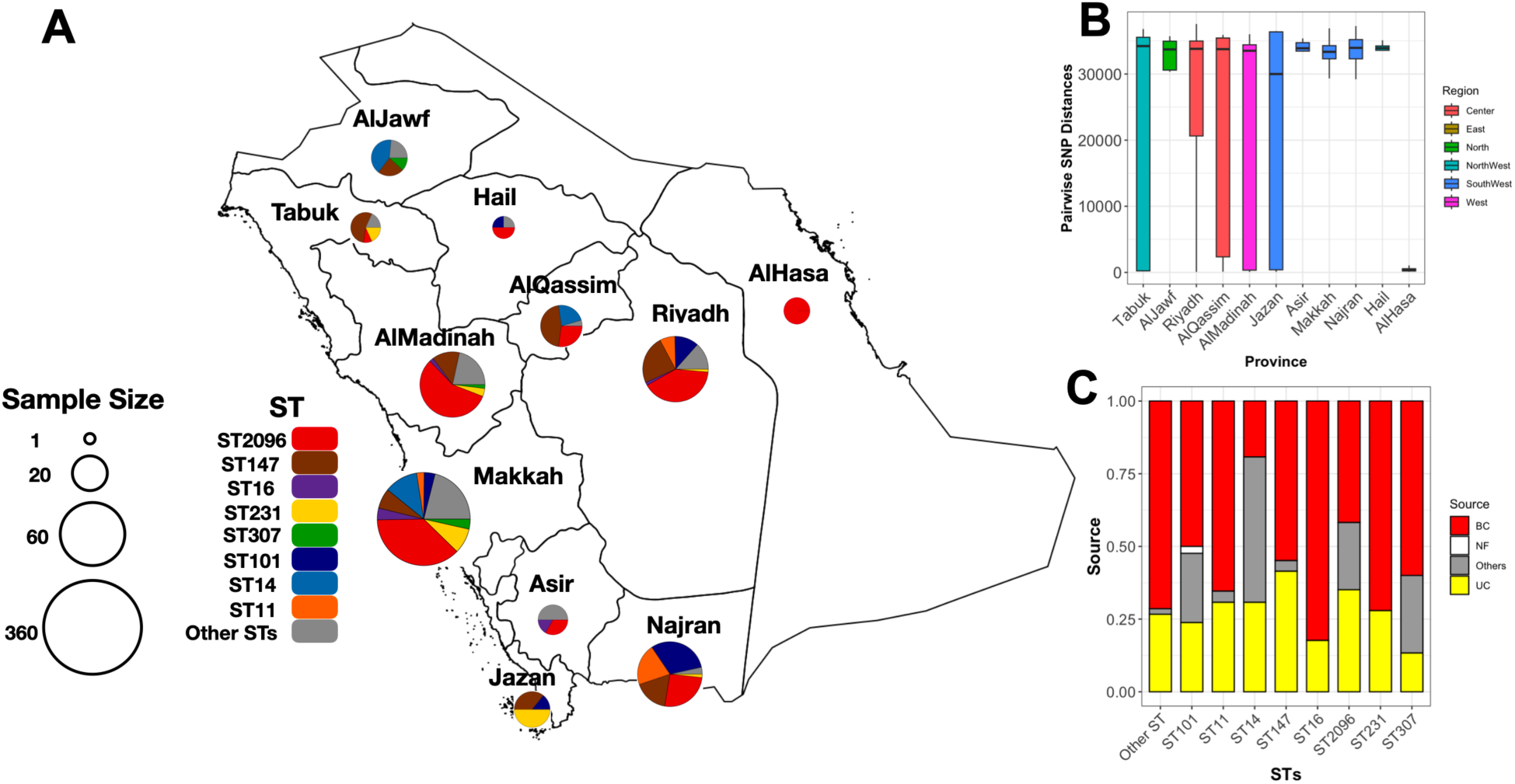
The distribution of *K. pneumoniae* clones in the study cohort. A) The frequency of STs across provinces. The size of the pie charts corresponds to the number of isolates. B) SNP diversity of isolates in each province. Diversity is measured as the pairwise SNP distance between isolates originating from the same province. The colours represent the regions in the country. C) Proportion of samples originating from different sources of isolation. The terms “BC” and “UC” stand for blood and urine culture, respectively.

**Figure 2.**
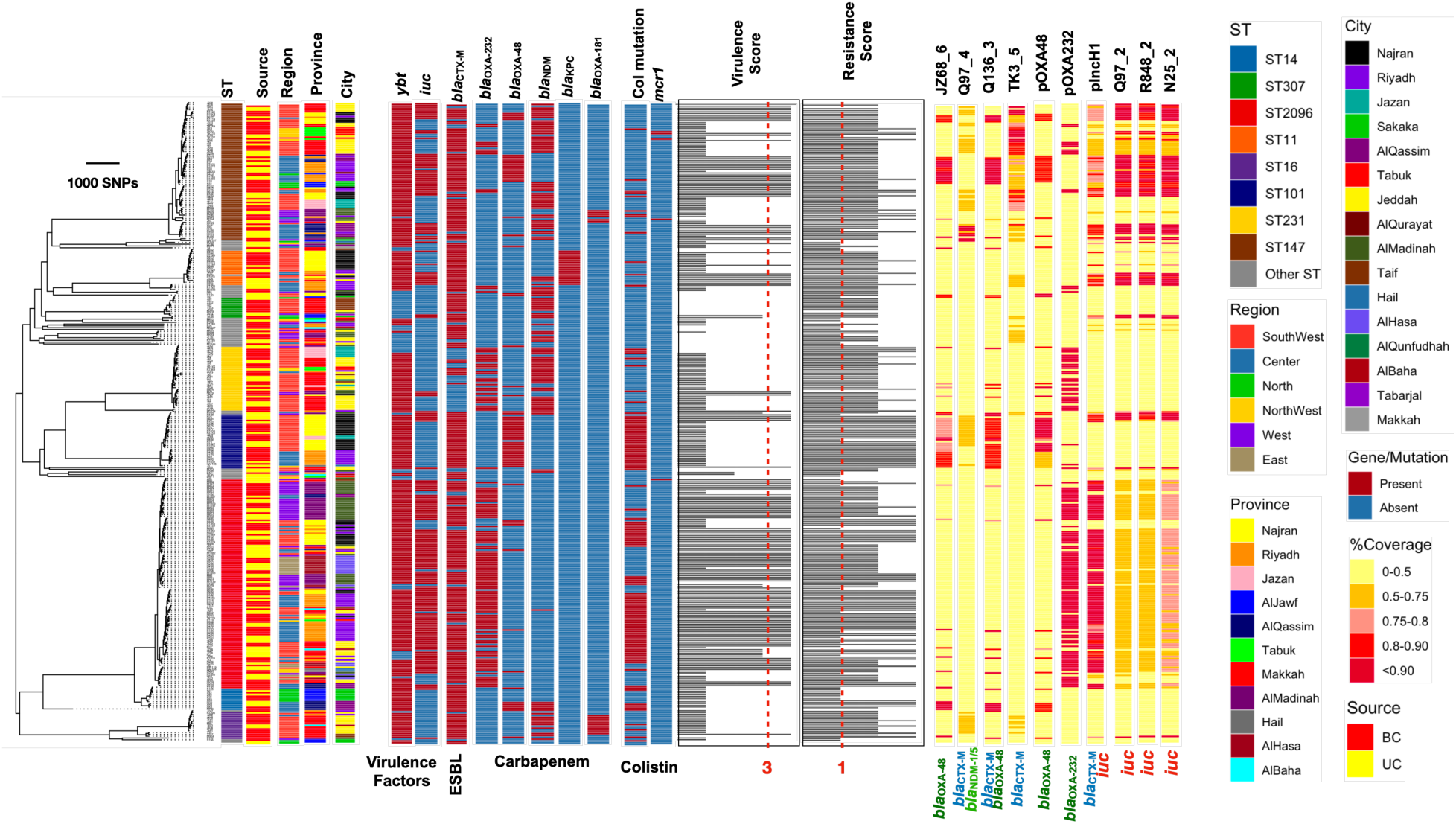
Phylogenetic tree for 352 *K. pneumoniae* strains recovered from hospitals in the current study and from a single-hospital study in Jeddah. The tree is an approximate maximum-likelihood phylogenetic tree from the alignment of 196,300 SNPs. Strains for STs with more than ten representative strains are shown. The resistance and virulence scores were computed in Kleborate. The dotted lines show the cut-off used for defining multidrug resistance and virulence factor genes. The plasmid bands correspond to the mapping coverage for the plasmids containing carbapenemase and ESBLs from long-read sequencing. “BC” and “UC” correspond to blood and urine cultures, respectively.

### Clonal expansion of multiple clones across the country

The distribution of major STs across the country revealed that dominant STs were spread across provinces. ST2096 was detected in 9/11 provinces, ST14 in 8/11 provinces, ST11 in 4 provinces, and ST231 in 5 provinces, mainly on the western side of the country (Figure 1A, Figure 2). All provinces contained multiple STs, except for the eastern province (AlHasa), in which all the genomes belonged to one clone, i.e., ST2096 (Figure 1A, Figure 2). Compared with those from other provinces, the samples from Riyadh and Makkah harboured more STs, i.e., 13 and 14 STs, respectively, which is likely because they are centres for religious tourism and global mixing. However, after accounting for the sample size, the diversity in the southern provinces, i.e., Jazan and Najran, was found to be greater than that in Makkah and Riyadh: For Jazan and Najran, the average pairwise SNP diversity was greater than the values for four and five provinces, respectively. Meanwhile, for Makkah and Riyadh, the average pairwise SNP diversity was greater than that in two and three provinces, respectively (p value from one-sided Wilcoxon signed rank test < 0.05) (Figure 1B). This shows that factors other than population size/density are involved in shaping the population diversity of the bacteria in the country. Moreover, the major clones appeared to have originated from various body sites, with no significant association with the body site of isolation, i.e., blood or urine (p-value from the proportion test > 0.05) (Figure 1C, Figure 2), indicating that all major clones occurring in the urine may cause bloodstream infections. Moreover, two of the nine samples recovered from food were ST101 and contained ESBL genes. The sharing of this ST with the clinical samples suggest potential epidemiological links between the food and human reservoirs.

### Mixing of Saudi samples with global and regional *K. pneumoniae* strains

We contextualized the collection of the isolates with the global collection and extracted epidemiological clusters (SNP clusters with a maximum SNP distance of 50). In total, eight clusters with samples from the collection under study and the global collection were extracted (Figure 3A). Clusters ST2096, ST437, ST147, and ST231 revealed the global dissemination of the clones with samples from four continents (Figure 3A). A closer examination of the population structure for clusters with at least ten Saudi strains revealed different levels of mixing of Saudi strains with the global population (Figure 3B): ST147 from Saudi Arabia forms a distinctive clade that mixes with and expands the clone in North America, specifically in Europe. In contrast, S2096, ST231, and ST11 showed five, three, three, and three incidences of introduction, respectively, from Saudi Arabia into other countries or from other countries into Saudi Arabia (Figure 3B). Mixing of the samples for these clones predominantly occurred with samples from the Middle East and South Asia, showing that Saudi clones constitute regionally circulating clones. All the global strains were of clinical origin.

**Figure 3.**
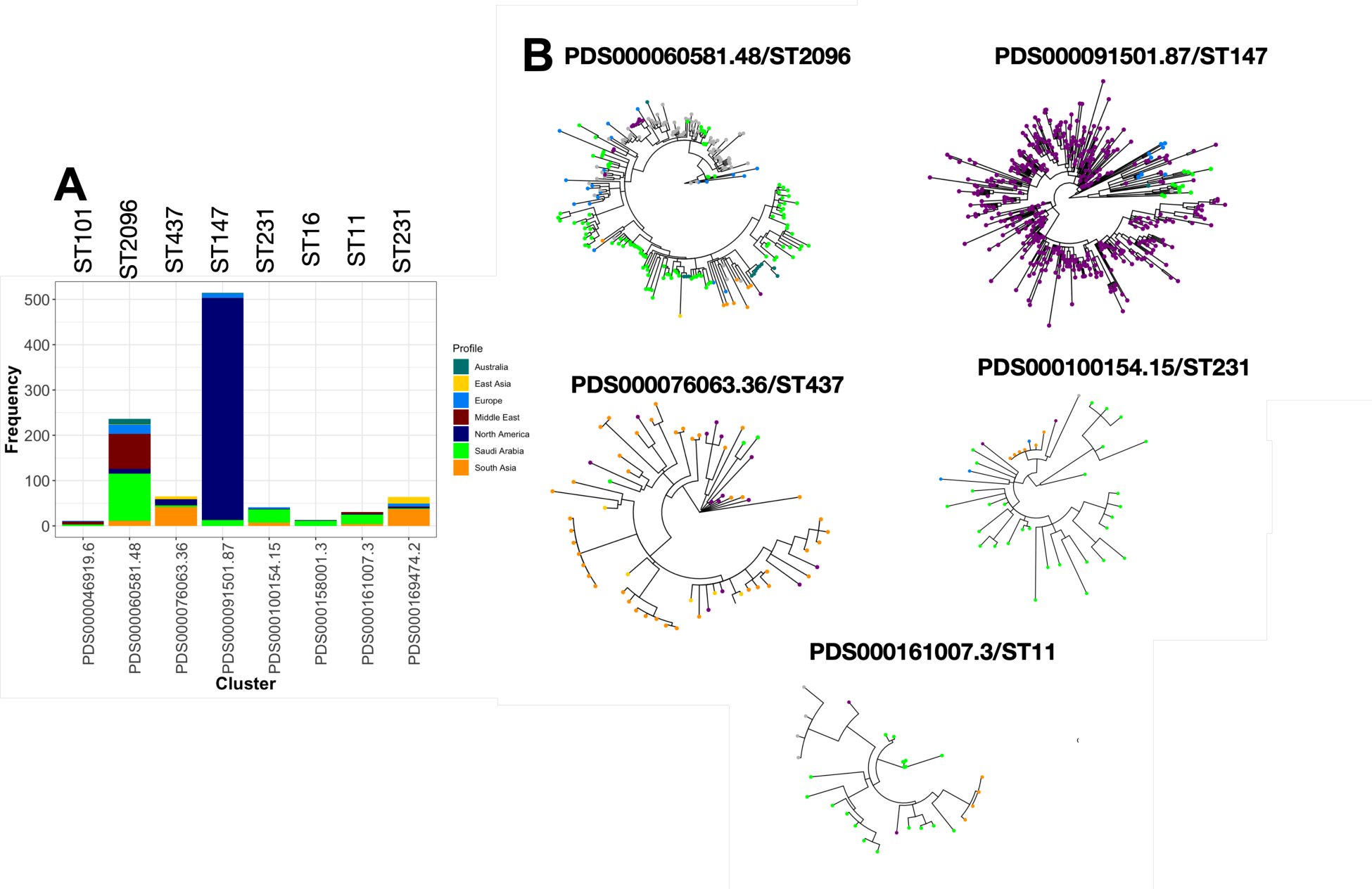
Mixing of Saudi clones with the global collection. A) The SNP clusters (clusters of genomes fewer than 50 SNPs apart) as derived from the pathogen detection database (see Methods for the definition of the clusters). B) Neighbour-joining tree for the clusters that contained more than two isolates from Saudi Arabia and the global collection in the database. The colour tips correspond to strains from Saudi Arabia and other regions. The PDS000158001.3 cluster included two isolates from the USA and eleven strains from Saudi Arabia and therefore is not shown. The PDS000169474.2 cluster included two Saudi strains and is therefore not shown.

### Higher resistance levels and frequencies of resistance determinants for dominant clones than for the rest of the population

The collection was extensively resistant to multiple classes of antimicrobial agents, including penicillin (95%), cephalosporin (83%), carbapenems (85%), aminoglycosides (67%), ciprofloxacin (84%), trimethoprim (82%) and nitrofurantoin (62%) (Figure S1). Except for the ST14 clone, which had a lower resistance level for aminoglycosides and cephalosporins than the other STs, the frequently detected ST clones had a higher resistance level than did the remaining population for β-lactams, aminoglycosides, and ciprofloxacin, which may account for their prevalence (Figure S1). This resistance phenotypic profile is congruent with the antimicrobial resistance determinants profile, in which major ST clones have a greater frequency of ESBL-positive strains, strains with non-ESBL β-lactamases, ciprofloxacin resistance mutations and carbapenemases than the remaining population (one-sided p value from proportion test < 0.05) (Figure S2A and S2B). In addition to a higher frequency of resistance genes, the carbapenemase genes across the major clones demonstrated diverse patterns across the phylogenetic tree, including the major clones (Figure 2): *bla*_NDM-1,_ *bla*_NDM-5_, *bla*_OXA-48_, *bla*_OXA-232_, *bla*_OXA-181_ and *bla*_KPC-2,_ which underlie carbapenem resistance in ST307/ST16/ST147/ST14, ST231/ST2096/ST101, ST16 and ST11, respectively. This variety of carbapenemase genes with reported prevalence on different continents points to the high population diversity of *K. pneumoniae* in the KSA and the admixture with global clones. In addition to carbapenem resistance, acquired resistance to colistin, another last resort antimicrobial, was detected in the ST2096 (*mcr1*), ST14 (*mcr1*), ST147 (*mcr1*) and ST261 (*mcr8*) clones. Colistin resistance mutations in the *mgrB(pmrB)* gene were found in 158 (3) genomes across the phylogenetic tree, with the highest prevalence in strains ST2096, ST14 and ST16 (Figure 2, S2B). These mutations were detected in all major STs, except for ST307, at a greater frequency than in the remaining population, indicating a link between the presence of the genes and prevalence.

### Strong link between resistance phenotypes and the presence of determinants for ESBLs, carbapenemase, aminoglycosides and ciprofloxacin

Integration of the genotypic data with phenotypic data indicated that the presence of carbapenem-acquired resistance genes accounted for an odds ratio of 10-20 across the β-lactams and an average resistance detection rate of 81% (Figure S3). The effect of ESBL on cephalosporins was smaller and was noticeable for cephalosporins cefepime (CRO) (odds ratio: 25) and cefolatin (CEF) (odds ratio 18) on average (Figure S3). The odds ratio for non-ESBL carbapenemase genes was smaller (average 4.5) and was significant for eight out of nine antimicrobial agents (95% confidence interval > 1). Mutations in genes encoding outer membrane proteins (Omp) had a significant effect on all β-lactams except carbapenems (Figure S3). The presence of β-lactam resistance determinants accounted for most of the total resistance (average 85%) for all determinants, except for mutations in the gene encoding SHV β-lactamases, which was, on average, 27% across antimicrobials (Figure S3). All resistant strains harboured aminoglycoside resistance genes, with an odds ratio of 18 for the resistance phenotype (Figure S3). While ciprofloxacin resistance mutations are linked with a high odds ratio of 25, accounting for 85% of resistance, the effect of acquired ciprofloxacin resistance genes was modest (odds ratio 1.01, RDR 45%) (Figure S3). Due to the high prevalence of resistance, many of the clades were linked to resistance, complicating GWAS analysis to pinpoint resistance biomarkers that are not linked to population structure (lineage effect). However, for some antimicrobials, we identified known/potential resistance determinants (p value for GWAS analysis < 0.05). This included the *bla*_NDM-1_ gene for imipenem, the multidrug transporter *emrE* gene for trimethoprim resistance, and a synonymous mutation at position 35 of the gene encoding an SDR family oxidoreductase, which might affect the expression level of the genes and confer tigecycline resistance, as reported for the Gram-negative *Acenitobacter baumannii* strains [36].

### *K. pneumoniae* population in the KSA contains multiple clones with high resistance and virulence levels

We further analysed the prevalence and phylogenetic distribution of key resistance and virulence genes in *K. pneumoniae* strains. In the collection of clinical *K. pneumoniae*, the frequencies of carbapenemase-, ESBL- and colistin-positive genes/mutations were 77%, 72% and 27%, respectively. These determinants were distributed across the phylogenetic tree (Figure 2), with major STs having a greater content of these key genes and hence a greater resistance score, which was significantly greater than that of the minor STs combined (p value from Wilcoxon signed rank test < 0.05) (see Methods) (Figure 4A). The highest resistance level was found for the ST101 clone, in which 82% of the genomes contained carbapenemase and colistin resistance determinants (Figure A). The patterns for key virulence factor genes were similar in that major clones had a higher virulence score than the remaining population (Figure 3B). All major STs, except for ST307, harboured the siderophore-encoding *ybt* gene. All strains belonging to ST231 and ST2096, the majority of ST16 strains, and three ST14 strains were of the *ybt14/ICEKp5* type. The ST11 and ST101 strains exclusively harboured *ybt 9/ICEKp3*, while all the ST16 and two ST147 strains contained the *ybt 10/ICEKp4*. The remaining ST147 strains harboured *ybt 16/ICEKp12*. The colibactin locus (*clb*) was identified in two ST294 strains and 36 strains from ST147, ST11, ST101, and ST16. Moreover, 163 strains contained the *iuc* locus, which included all major STs, except for ST307, among which 3 strains from ST231 contained the *iuc5* locus and the rest contained the *iuc1* locus (Figure 4B). The integration of resistance and virulence profiles for the strains sequenced for this study resulted in the identification of strains from ST101, ST11, ST147, ST16, ST2096, ST231, ST307, ST751, ST45 and ST383 as having strains with a dual resistance/virulence risk (MDR-hv), i.e., resistance score ≥1 and virulence score ≥3. ST751 and ST383 each had one representative genome, were also identified as MDR-hv (Figure 2). The largest MDR-hv group was ST2096, reported previously from a single-hospital study in Jeddah [16]. The ST2096 genome mostly contained the *iuc* locus and carbapenemase genes (*bla*_OXA-48,_ *bla*_OXA-232_ and *bla*_NDM-1_) and has recently expanded (Figure 2, 4C). The single ST751 strain was obtained from blood and contained *iuc*, *bla*_KPC-2_ and *bla*_CTX-M-65_. The ST383 strains were also from blood and, in addition to the *iuc* locus, contained *bla*_CTX-M-14_ and dual carbapenemase *bla*_NDM-1_ and *bla*_OXA-48_ genes.

**Figure 4.**
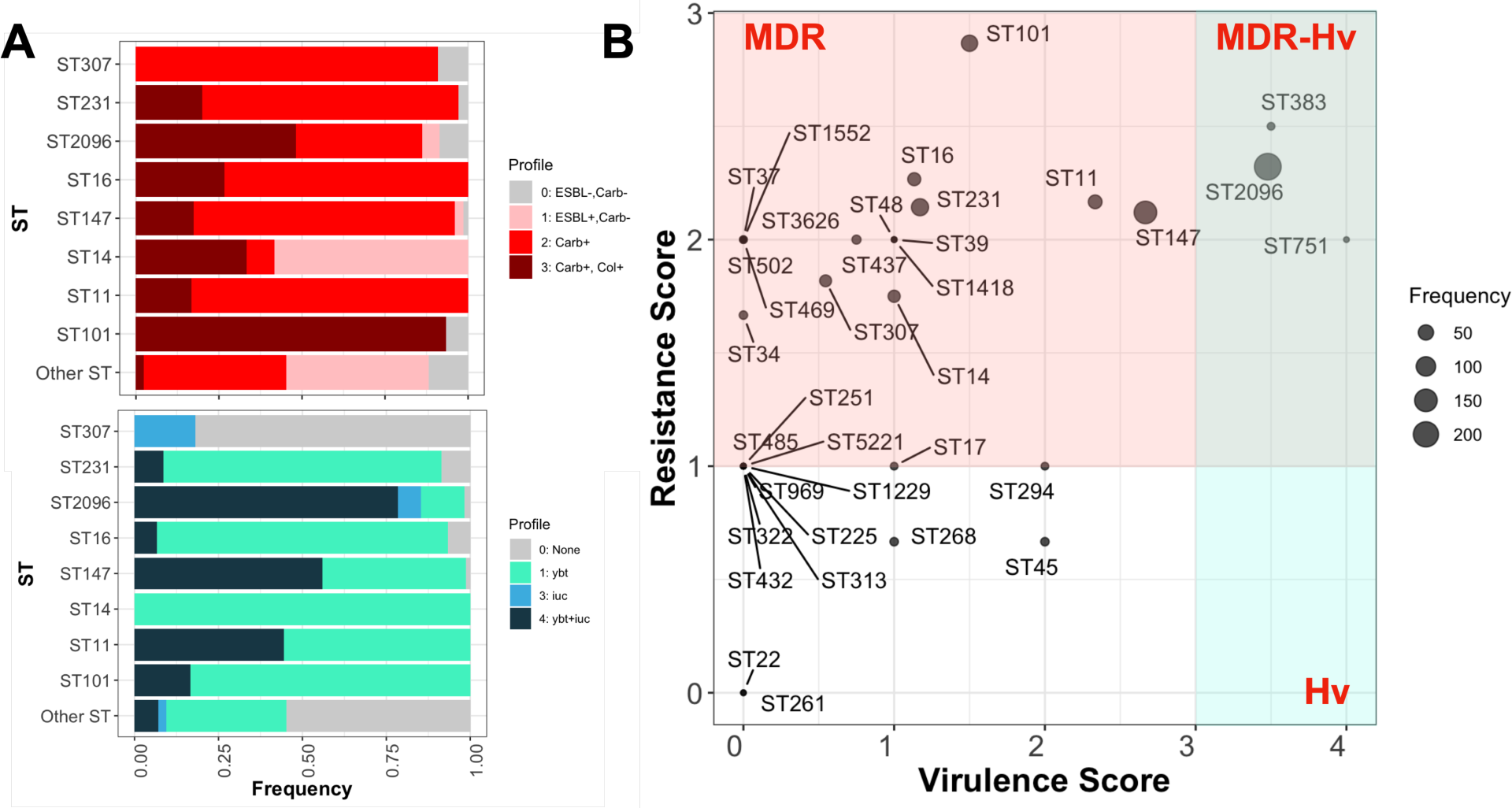
Frequency of clinically significant resistance and virulence genes across major clones of *K. pneumoniae* from blood stream infections in the KSA. A) The frequency of ESBL, colistin and carbapenemase genes across STs. The numbers show the resistance scores (see methods for more information). B) The distribution of aerobactin (*iuc*) and yersiniabactin (*ybt*) across major clones (STs) and other STs. The numbers show the virulence scores (see methods for more information). C) The integration of the virulence score and resistance score for the STs in the population. The shaded areas define the hypervirulent, multidrug resistant and hypervirulent-MDR resistant space. The scores and definitions of hypervirulence and MDR were adopted from Kleborate, and details are provided in Supplemental Table S1. The x- and y-axes correspond to the average virulence and resistance scores, respectively.

### Higher death rates for the ST231, ST16 and ST11 strains and strains with carbapenemase and yersiniabactin genes

The availability of mortality data for bloodstream infections allowed us to determine the mortality rate associated with infections caused by different STs and by strains with different patterns of ESBL, carbapenemase, and virulence genes. In total, the mortality and sepsis rates for the blood culture samples were 0.54 and 0.18, respectively, where 33% of deaths were due to sepsis. The pooled death rates were 14% at 7 days, 22% at 14 days, 34% at 30 days and 39% at 90 days, which are comparable to the reported rates from a meta-analysis of *K. pneumoniae* bacteraemia [37]. The pooled mortality odds ratios (ORs) of strains with carbapenemase-encoding genes versus strains without these genes were 1.59 (95% CI 1.18-8.76) at 7 days, 1.73 (95% CI 4.31-7.42) at 14 days, and 1.42 (95% CI 3.01-3.49) at 30 days. These values were lower than those reported in previous studies [37] and were not significant. This might have occurred because the strains with no carbapenemases harboured other virulence/resistance genes, which made them as fatal as the strains with carbapenemases in our collection. The highest mortality rates were found for strains ST11 (0.76), ST16 (0.76), and ST231 (0.71), which were significantly greater than the rates for the remaining population (p value from the proportion test < 0.05) (Figure S4A). In the CC14 clade (ST2096 and ST14), the mortality and sepsis rates for ST2096 were significantly greater than those for the ST14 clones (p value from one-sided proportion test < 0.01), which reflects the higher virulence and resistance score for the ST2096 strains than for the ST14 strains, consistent with the findings of a single-hospital study in Jeddah (Figure S4A) [16]. The lowest mortality rates were observed for ST14 (0.14) and ST307 (0) (Figure S5). For patients with reported sepsis, ST231 infection had the strongest association with sepsis (p value from the proportion test < 0.01) (Figure S4B). The integration of key virulence and resistance genes revealed a greater death rate for samples harbouring *ybt* and samples with the *iuc* and *ybt* loci; in the former case, the rate was significantly greater than that of the remaining population (Figure S4C). Among the resistance genes, the presence of carbapenemase genes led to a significantly greater rate, than the rest of the population (Figure S4C). Our findings highlight the clinical manifestations of resistant and virulent clones, although the scarcity of strains with certain virulence and resistance gene profiles (e.g., only three samples harbouring *iuc* without the *ybt* locus) prevented us from accurately estimating the mortality effects of all combinations of virulence and resistance genes.

### Sharing of plasmids encoding virulence and resistance genes between major clades

We next investigated the resistance-linked plasmid of the population through an integrated third and second sequencing approach on selected strains (see Methods). In total, 29 plasmid fragments encoding resistance genes were retrieved from 20 isolates selected for third-generation sequencing (see Methods). These plasmids were combined with three resistance plasmids from ST2096, i.e., pNDM, pOXA232 and pOXA48, which were isolated from the ST2096 strain from a single-hospital study in Jeddah [16] (Figure 5). Among the 32 plasmids, 19 plasmids occurred in more than one clone, as indicated by higher coverage, i.e., > 90% of the plasmid backbone when used as a reference genome (Figure 5). Three plasmids, R848_3, Q97_4, and Q136_3, which were recovered from strains ST11, ST147 and ST1418, respectively, contained both the ESBL and carbapenemase (*bla*_KPC-2_ and *bla*_OXA-48_) genes. Four resistance-linked plasmids from ST2096, ST101, ST14, ST11 and ST147 also contained the hypervirulence gene *iuc* (Figure 5). Carbapenemase genes were found on diverse plasmid backgrounds: the *bla*_OXA-48_ gene was found on ∼100 kb plasmids both alone (pOXA48) and in conjugation with *bla*_CTX-M-15_ (Q136_3) on plasmids from the CC14 clone, i.e., ST2096 and ST1418 (Figure 4). Multiple clades included the single epidemic pOXA48 plasmid, which was recognized for its rapid dissemination across lineages and hospitals [38]. Similar to the *bla*_OXA-48_ gene, the *bla*_NDM-1_ and *bla*_NDM-5_ genes were also found in combination with other resistance genes on plasmids 100-130 kb in size from the ST147 and ST307 strains, in which the plasmid also contained the ESBL gene. The *bla*_KPC-2_ gene was located on a plasmid containing the ESBL gene in ST11 strains. In contrast to *bla*_OXA-48_, *bla*_NDM-5_, and *bla*_NDM-1_, the *bla*_OXA-232_ gene was found on small plasmids identified in the ST2096 isolates, as reported previously [16] and appears to have been present across the phylogenetic tree in ST14, ST147 and ST231 (Figure 2). The *bla*_OXA-181_ was found on a plasmid with other resistance genes, mainly in the ST16 strains. These results point not only to extensive sharing of resistance plasmids across major clones, showing potential horizontal transfer in the population, but also to the variety of plasmid contexts for the carbapenemase genes.

**Figure 5.**
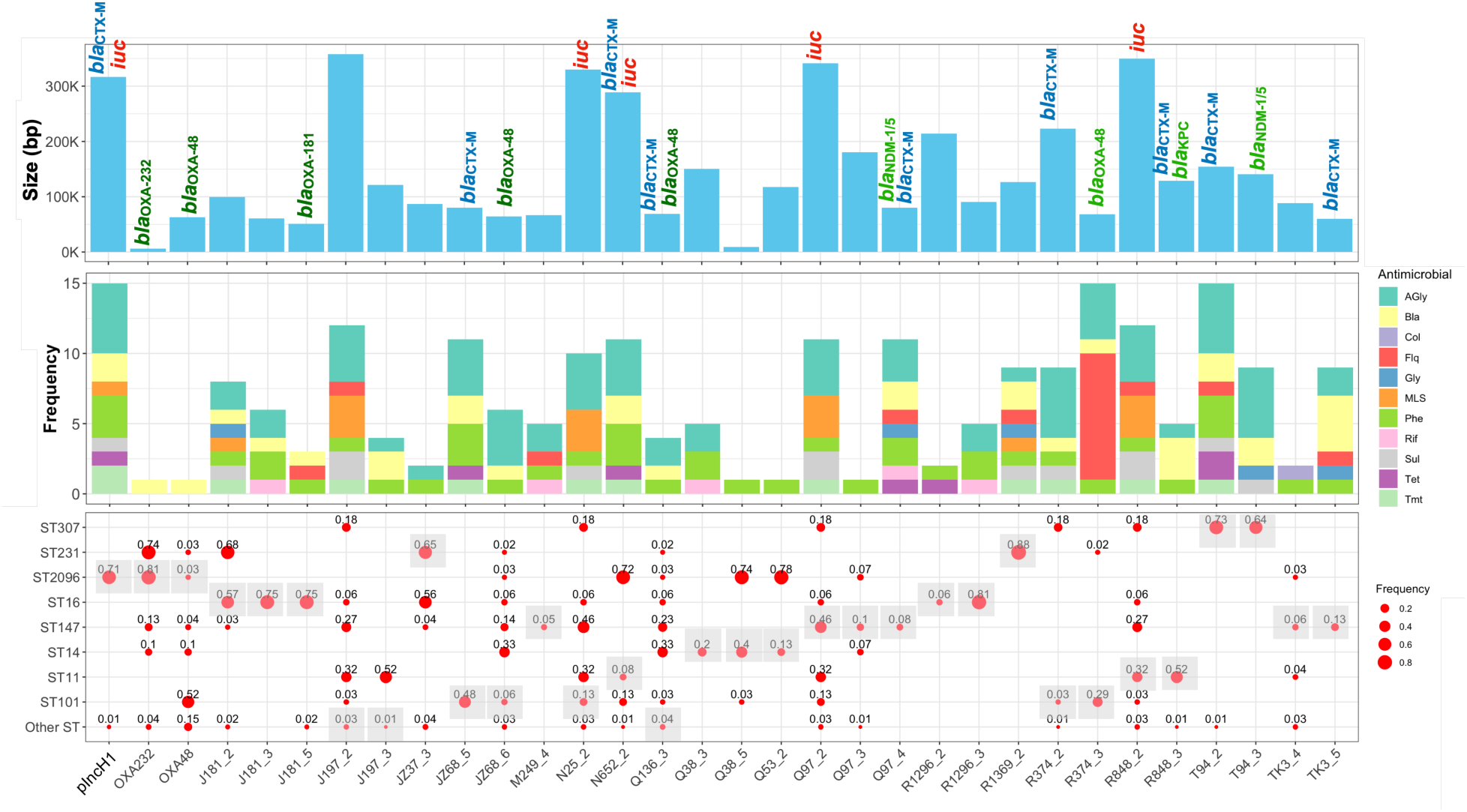
The retrieved plasmid fragments that contained a resistance gene. The top panel shows the length of the plasmid in base pairs. The bubble plot shows the frequency of plasmids across major clones. We mapped the short reads to the plasmid fragments and measured the mapping coverage. The shaded boxes show the STs of the strains from which the plasmid fragment was retrieved.

### Configurations of resistance virulence genes on the same plasmid backbones

We characterized the genomic architecture of seven mosaic plasmids with ESBL and carbapenemase genes and those with virulence and resistance genes (Figure S5). Both plasmids JZ68_6 (from an ST101 strain) and Q136_3 (from an ST1418 strain) resembled the pMpOXA-48 plasmid and contained two islands for *bla*_OXA-48_ and a resistance cassette; however, the resistance cassette in Q136_3 had a *bla*_CTX-M-27_ gene instead of an aminoglycoside phosphorylase gene. Both plasmids had a broad phylogenetic distribution and contained *Tra* genes encoding conjugation pilus (Figure S6, 2). The Q97_4 and TK3_5 plasmids were of the pFIBK and pR types, had a narrow phylogenetic distribution and were limited to the ST147 clade (Figure 5). Both plasmids contained two resistance cassettes, one of which contained the carbapenemase genes *bla*_NDM-1_ for Q97_4 and *bla*_NDM-5_ for TK3_5, and the other contained ESBL. In contrast to the pMpOXA-48 plasmids, both plasmids appeared nonconjugated because they lacked a full set of *Tra* genes (Figure S5). The mosaic virulence plasmids N25_2 (from the ST101 strain), Q97_2 (from the ST147 strain) and R848_2 (from the ST11 strain) had a broad phylogenetic distribution and occurred in at least three major ST groups. Like the pNDM plasmids, which were characterized previously in ST2096 strains and accounted for the convergence of resistance and virulence in these strains [16], all plasmids were ∼300 Kbp in size and included Tra genes (conjugation genes), a virulence cassette with the *iuc* locus and *iutA* gene, and a resistance cassette, which varied across the plasmids (Figure S5). The virulence cassette was flanked by copies of the *rmpA* and *rmpA2* virulence genes, i.e., regulators of the hypervirulent mucoid phenotype, which were present only in Q97_2 and R848_2 and not in N25_2 (Figure S5). The variation in the presence of resistance and virulence genes on the backbone of highly similar plasmids in different STs suggests the dynamics of the virulence and resistance cassette during evolution in clinical settings.

### ST231, ST147 and ST2096 have recently expanded across the country and show signatures of between-hospital and between-cite transmissions

The coexistence of the clones motivated us to examine the dynamics of the major clones in the population (Figure 6). The temporal signals in the genomes of five main clones, ST101, ST11, ST2096 and ST147, allowed us to reconstruct population dynamics and examine population growth. The clones had a comparable clock rate (Figure 6A). ST2096, ST147 and S231 clones formed in 2004 (95% CI: 2001, 2007), 1992 (95% CI: 1964, 2002) and 1998 (95% CI: 1981, 2014), respectively, which were more recent than those of the ST101 and ST14 clones (Figure 5A). The clone ages for ST2096 and ST147 were close to the reported clone ages in other studies, suggesting that the population diversity of these clones in Saudi Arabia reflects global diversity (Figure 5A) [16, 39]. The skyline growth plot for the clones indicated rapid initial population expansion over a period of 20 to 30 years for ST2096, ST147 and ST231 (Figure 6B). For ST2096 and ST147, the population stabilized after the initial increase, while for ST231, after a decrease, the population grew again (Figure 5B). The average migration rates of 0.0204 (95% CI: 0.0125, 0.0296), 0.0224 (95% CI: 0.0111, 0.0369), and 0.031 (95% CI: 0.00498, 0.0708) strains per year for ST2096, ST147 and S231, respectively, were greater than those for for the ST101 and ST14 clones (Figure 6A). Rapid dissemination of these clones is inferred to have occurred across the country over 7 years and included cities in the western, northern and central regions of the KSA (Figure 6C). For all STs, a frequency of approximately 1-2% of the potential transmissions, i.e., genomes with fewer than 10 SNPs apart, was found (Figure 6D) (see Methods). For ST14, transmissions exclusively occurred within the same hospital, while for ST101, half of the transmissions were for strains from different hospitals within the same city. Moreover, consistent with the expansion pattern from the phylogeographic analysis, highly similar pairs of genomes were detected for the ST2096, ST147, and ST231 strains across various cities (Figure 6D). This similarity was observed not only among strains originating from different hospitals within the same city but also among those from different cities, pointing to the rapid spread of these clones throughout the country on an epidemiological time scale.

**Figure 6.**
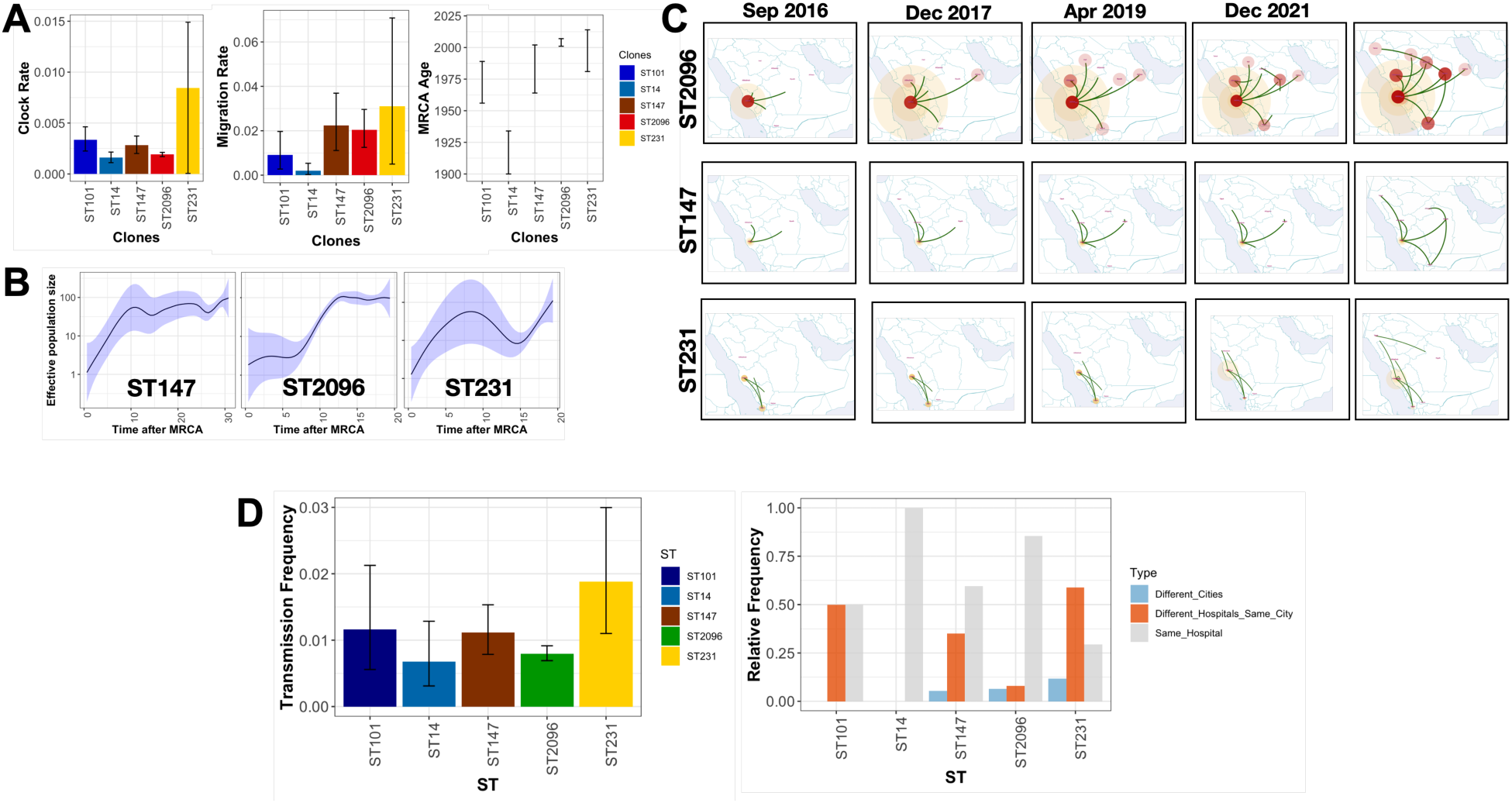
Population dynamics of major clones. A) The phylodynamic parameters from the phytogeography analysis. Clock rates show the rates based on the number of substitutions per site per year. The migration rates show the parameters for all the cities and the rates. The unit for migration is the number of migration events per year. The most recent common ancestor (MRCA) age indicates the time at which the clone formed. The error bars show the 95% confidence intervals. B) The skyline growth plot. The shaded plot shows the 95% confidence intervals. C) The recent spread of ST147 and ST2096 across the country. The trajectories show the inferred dissemination (migration routes) across the country. The circle sizes and colour intensity and rate show the relative marginal rate per city. D) The transmission frequency shows the frequency of transmission events, defined as the pairwise genomic distance of less than 10 SNPs divided by all possible transmission events. The error bars show the 95% confidence intervals from the binomial test. The relative frequency shows the distribution of transmissions within the same hospital, between hospitals within the same city or between cities.

### Concurrent expansion of ST2096, ST147 and ST231 and the acquisition of resistance and virulence plasmids

We investigated the dynamics of major clones, antimicrobial resistance, virulence, and associated plasmids. ST2096, the largest clade with nationwide expansion, is a dual-risk clone due to a mosaic plasmid containing both resistance and virulence genes (pIncH11). The clade also harbours the carbapenemase encoding pOXA48 and pOXA232 plasmids, which are present in distinctive clades (Figure 7). One ST2096 strain also acquired *bla*_NDM-5,_ showing the ongoing evolution of carbapenem resistance in the clade (Figure 7). ST147 is a more long-standing clone than ST2096 and consists of three subclades that have expanded over the past ten years and contain different variants of the yersiniabactin gene, which suggests multiple independent introductions of ST147 sublineages into the country (Figure 7). The inferred age of the ST147 clone concurs with its reported emergence time in the early 1990s and the expansion of distinctive clades with different patterns of resistance and virulence genes [6]. One clade that carries *ybt16* is limited to the north, whereas the clade with the *ybt9* gene shows a nationwide distribution across six regions and twelve hospitals (Figure 7). The clade with the *ybt16* gene also had GyrA-83Y and GyrA-87A mutations, while the clade with *ybt9* had GyrA-83I mutations (Supplemental Table S1), showing the divergent evolutionary history of ciprofloxacin resistance in the two clades, driven by ciprofloxacin treatment [6]. The lineages in both clades carried virulence genes, i.e., the *iuc* locus and *rmp* on the Q97_2 plasmid, as well as multiple plasmid-borne carbapenemase genes, *bla*_OXA-48_, *bla*_OXA-181_, *bla*_NDM-1_, *bla*_NDM-5_ and *bla*_KPC-2,_ independently (Figure 7). The acquisition of plasmids identical to the Q136_6 plasmid, which also carried ESBL, TK3_5 and TK3_5, supported carbapenemase resistance in the expanding ST147 clade with *ybt9.* As in ST2096, the ST231 clade consisted of a clade expanding across the southern, western and central regions of the KSA (Figure S6). Three isolates lost the *ybt* clade, and none of the strains contained the *iuc* locus, making the clade less virulent than other major clones (Figure S6). However, 76% of the strains contained both the NDM and OXA232 carbapenem genes, the latter of which was carried on the OXA232 plasmid shared with ST2096. ST101 consisted of two recent clades with limited geographical spread. One clade included strains with an ESBL *bla*_CTX-M-15_ encoding plasmid (JZ68_5), which is exclusive to the ST101 clade (Figure S6). Some strain in the same clade gained a second copy of ESBL *bla*_CTX-M-15_ situated on a mosaic ESBL/virulence plasmid (N652_2), similar to ST2096 (Figure S6). The prevalence of *bla*_OXA-48_ was due to the presence of the pOXA48 plasmid, which is shared with ST2096, and JZ68_6 plasmid, which is similar to pOXA48, and the R374_3 plasmid, which contains resistance genes other than OXA48 (Figure 4, S6). The ST14 clade is similar to the ST147 clade in that it contains two recently expanding clones with distinctive *ybt* gene types (Figure S6). One clade that mainly expanded in the west but was detected in the north contained *bla*_NDM-1_ and *bla*_OXA-48_ genes, which are likely to be located on the ESBL/carbapenemase plasmid (Q136_3) from another CC14 ST1418 strain (Figure S6). Taken together, these findings demonstrate that the evolution of antimicrobial resistance through the acquisition of plasmids occurred on the same timescale as clonal expansion.

**Figure 7.**
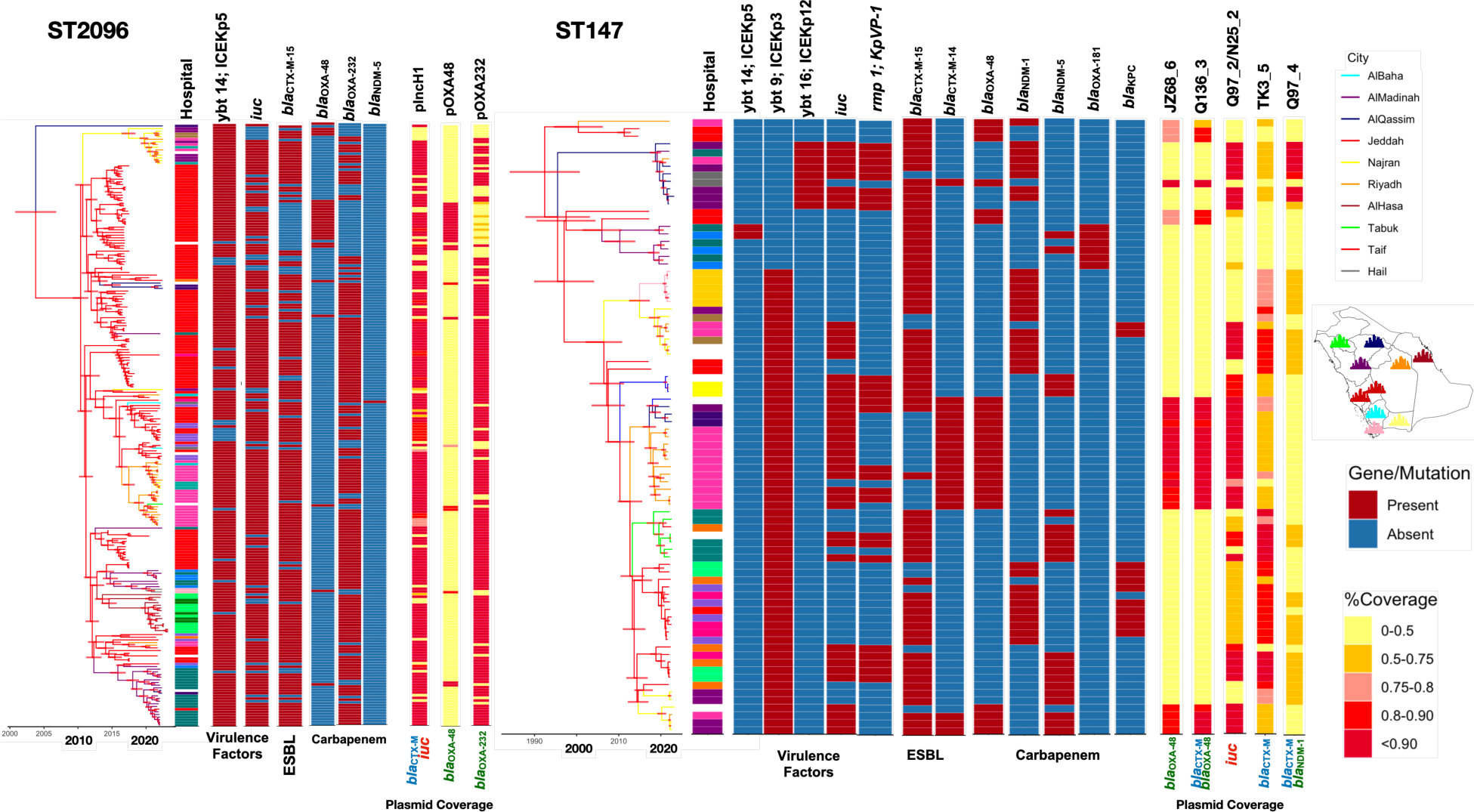
Phylogenetic trees for ST2096 and ST147. The trees are Bayesian trees reconstructed by discrete state simulation. Horizontal red lines correspond to the 95% highest posterior density (HPD) interval. The colours correspond to the inferred ancestral states of the cities. The genes were identified in Kleborate. The plasmid strips show the mapping coverage of short reads against the plasmid reference genomic fragments.

## Discussion

### Summary of results

This study presents a large-scale genomic analysis of the epidemiology and evolution of multidrug-resistant *K. pneumoniae* strains in the Kingdom of Saudi Arabia. Our systematic collection included a snapshot of clinical isolates from hospitalized patients for a large hospital network across the KSA. Our results indicate the concurrent expansion of multiple clones across hospitals, with the dominance of a few clones, most notably ST2096, ST147, and ST14, which commonly harbour plasmids with multiple carbapenemase, ESBL, and virulence factor genes. We also found independent introductions and subsequent expansions of clones representing sublineages of the dominant STs into the country. Although previous studies reported the prevalence of some of these STs across the Middle East and GCC [12, 16, 35], no major study has revealed the dynamics of the resistance clones and plasmids and virulence at high resolution within a consistent framework.

### Population diversity in the KSA shows clonal expansion of global and regional MDR/hvKp clones

The evolution of multidrug-resistant *K. pneumoniae* is characterized by the expansion of clones at the hospital level driven by nosocomial transmission routes, followed by regional and global expansion due to strong selective pressure from antimicrobial agents. The local epidemiological and clinical characteristics of *K. pneumoniae* have been reported in multicentre studies in different countries [6, 33, 40–42], revealing expansions of both endemic clones and imported clones, with clonal expansion mostly caused by the globally known ST258 lineage. Our results concur with the findings of these studies, showing similar clonal expansion of globally and regionally circulating clones in the KSA. In addition to clonal expansion, our integrated high-resolution analysis of both chromosome and plasmid sequences highlighted not only the diverse backgrounds of carbapenemase genes but also concurrent plasmid expansion across divergent lineages. This sharing of plasmids may account for the concurrent rapid expansion of these clones. Consistent with our findings, carbapenemase-carrying plasmids have been shown to occur in diverse lineages and are associated with previously reported high-risk clones that frequently spread across healthcare networks [43]. In addition to the global high-risk clones, our results highlight the significance of ST2096, which appears to be currently limited to Saudi Arabia, the Middle East, and South Asia [16, 44–46]. An expanding ST2096 clone was reported in a single-hospital study over an 8-year period in Jeddah and was proposed to be an emerging clone, with pervasive transmission within the hospital wards and links to sepsis and adverse clinical manifestations [16]. Therefore, we underscore the carbapenem-resistant ST2096 as an emerging MDR/hvKp-endemic clone, which, similar to ST258, which emerged from ST11 and rapidly spread worldwide, descended from the sister CC14 clade of ST14 and is becoming an endemic clone. The success of ST2096 clone, compared with that of ancestral ST14, is due to enhanced pathogenicity and resistance, which is driven by the coacquisition of plasmids carrying MDR/hvKp and carbapenemase genes [2].

### Limitations

The emergence of multidrug-resistant *K. pneumoniae* strains has been reported in clinical [8], urban (9), and agricultural settings (10) and is a worldwide phenomenon [11–13]. To fully characterize the reservoirs and transmission dynamics of these endemic clones, unbiased and comprehensive surveillance is warranted. Despite the depth and breadth of our sampling, several limitations should be noted. First, our collection included only a few environmental samples. Epidemiological links with environmental sources have been reported for *K. pneumoniae* strains in a previous study [47], suggesting spill-over of samples from hospitals into linked environmental sites [48]. Although the sample size from food was small in our study, we reported the sharing of ESBL-harbouring ST101 strains between humans and food. This finding points to epidemiological transmission routes, which may become increasingly detectable with broader and more in-depth sampling. Furthermore, our collection predominantly consisted of pathogenic and multidrug-resistant strains and therefore did not include nonresistant or nonpathogenic strains. These strains might become pathogenic and multidrug resistant through plasmid acquisition. Finally, our study covered one and a half years and therefore did not reveal long-term clonal dynamics. In future studies, sampling could be extended to include not only environmental strains but also susceptible nonpathogenic strains in a purpose-designed and unified sampling framework over an extended period to fully reveal the details of *K. pneumoniae* transmission routes in the KSA.

### Conclusion

This study highlights a major need for establishing a nationwide program critical for informing national and regional infection prevention and surveillance efforts. We demonstrated that precision epidemiological analysis through whole-genome sequencing offers an effective way to elucidate transmission dynamics at the regional level. With the extension of the study to cover collections from geographically widespread healthcare centres across the Middle East and North Africa (MENA) regions, which are currently significantly underrepresented in genomic databases, the approach may yield further significant insights over a wide range of geographical scales. Understanding the prevalence and epidemiological characteristics of *K. pneumoniae* infections in the MENA region will help in the design of interventional strategies tailored to the region.

## Data Availability

The genomic data have been deposited in the European Nucleotide Archive (ENA) under the study accession number PRJEB66182. Moreover, the assemblies have been uploaded to the NCBI GenBank database under the accession number PRJNA1018815. Detailed metadata associated with the genomes are provided in Supplemental Table S1.

## Acknowledgment

DM, MM, JH, DA, UA, and GZ were supported by KAUST faculty baseline fund (BAS/1/1108-01-01). AP is supported by KAUST baseline (BAS/1/1020-01-01). The authors extend their appreciation to the Deputyship for Research and Innovation, “Ministry of Education” in Saudi Arabia for funding this research (IFKSUOR3-478). We would like to thank Liliane Okdah for the help with the experimental protocols.

## Author Contribution

J.H. and A.Y.A. contributed equally to this work. J.H. and A.Y.A., U.H.A., M.M. and G.Z. conducted the experimental and computational research. D.A., M.B., A.A.A., A.A.R., S.B., D.B., A.N.A., Z.A.A., S.M.A., P.-Y.H., M.A., and W.A.S. provided resources. A.P., W.A.S. and D.M. conceived and supervised the study. All authors have read and approved the manuscript.

## Supplementary Figures

**Figure S1.**
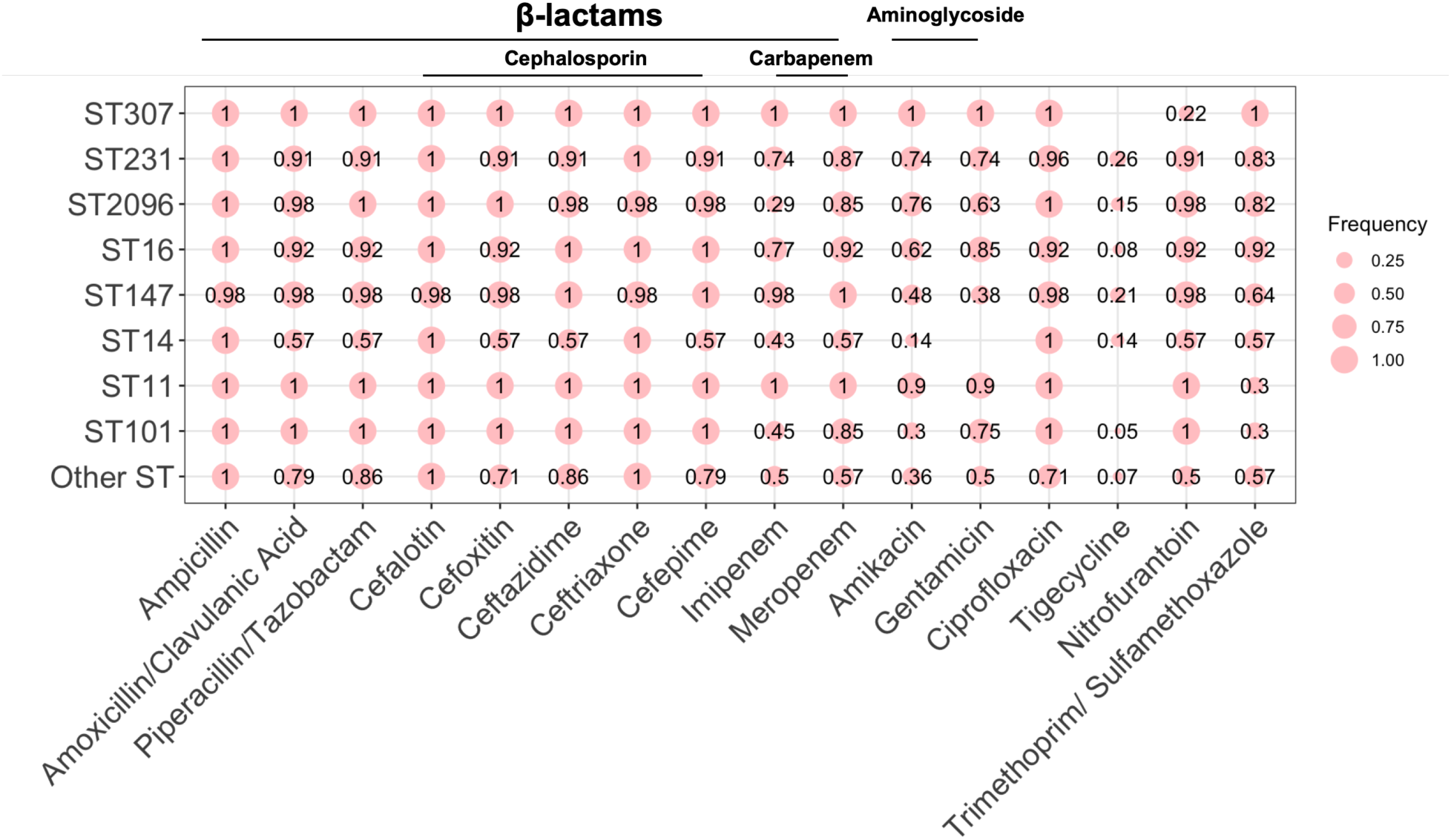
The frequency of resistant strains in the collection under study. The numbers show the relative frequencies. The circle sizes correspond to the frequency.

**Figure S2.**
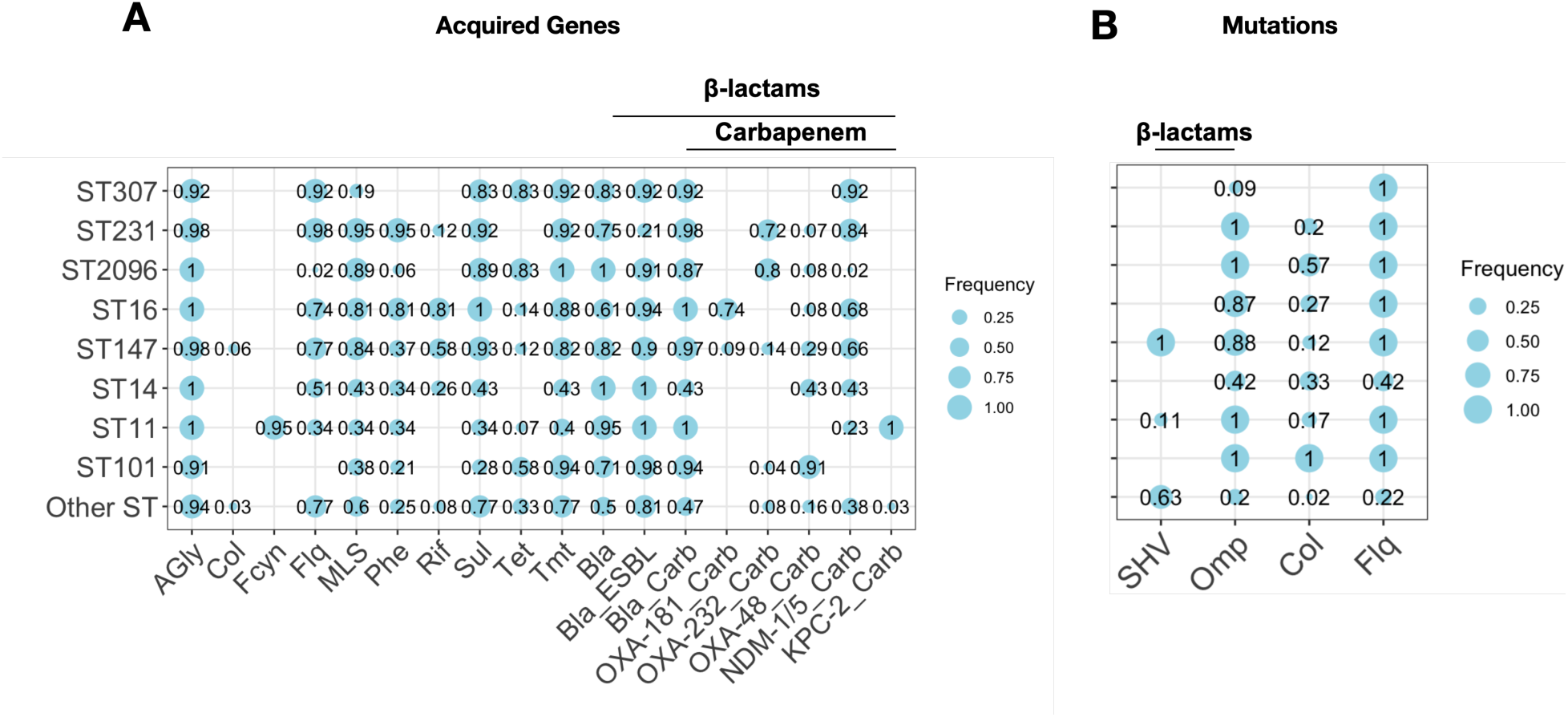
The frequency of acquired resistance genes and mutations across STs with more than 1% frequency in the collection under study. The numbers show the relative frequencies. The circle sizes correspond to the frequency. The resistance profile shows the distribution of major resistance determinants, i.e., Carb (carbapenemase), ESBL, AGly (aminoglycosides), Flq (fluroquinolone) and Tet (tetracyclines) (see Supplemental Table S1 for the full list of resistance determinants).

**Figure S3.**
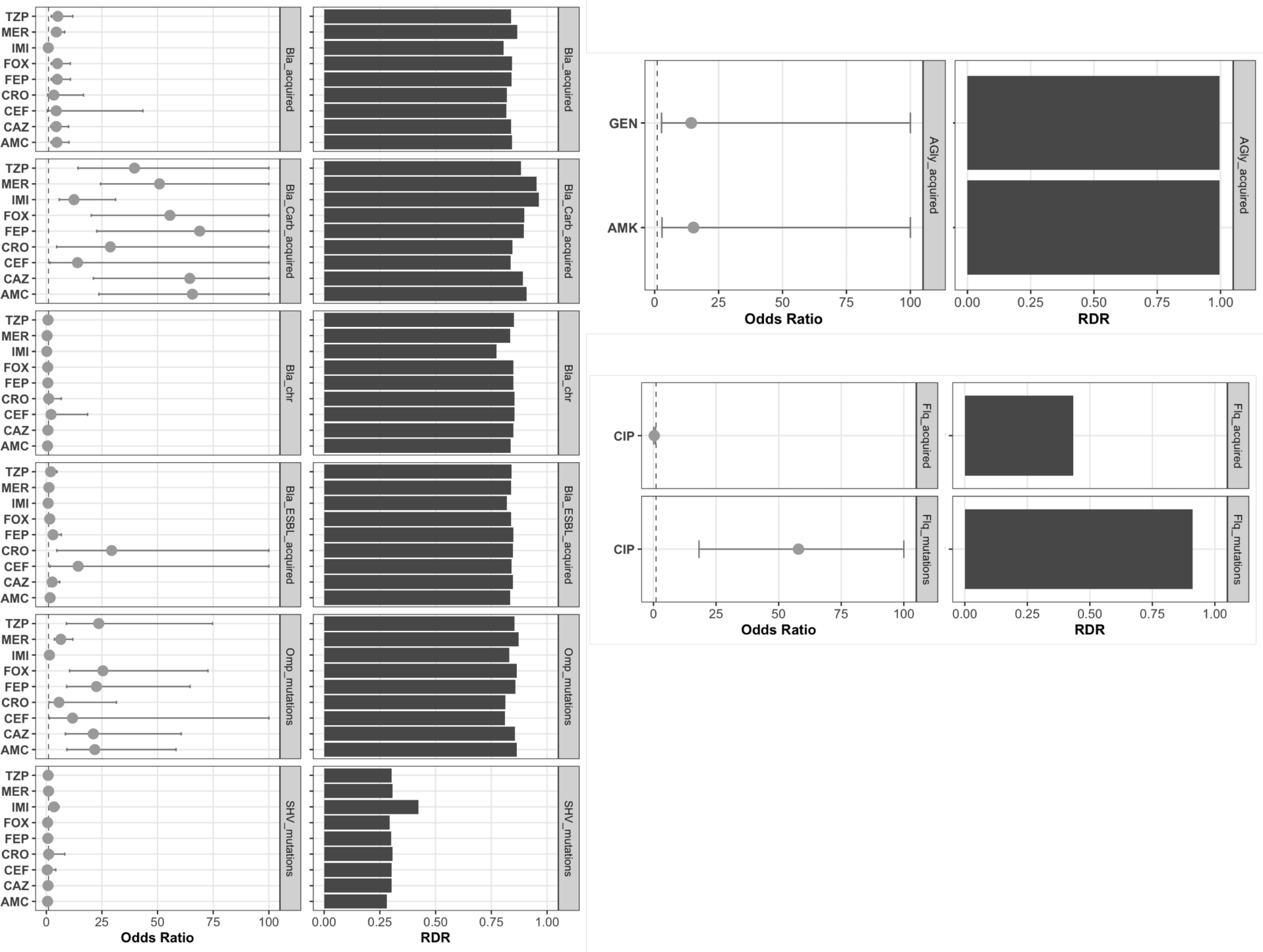
Integration of phenotype–genotype data. The odds ratio shows the odds of resistance for the presence of antimicrobial resistance determinants detected in Kleborate. RDR stands for the resistance detection rate, which corresponds to the ratio of resistance genomes with the resistance determinant divided by the total number of genomes.

**Figure S4.**
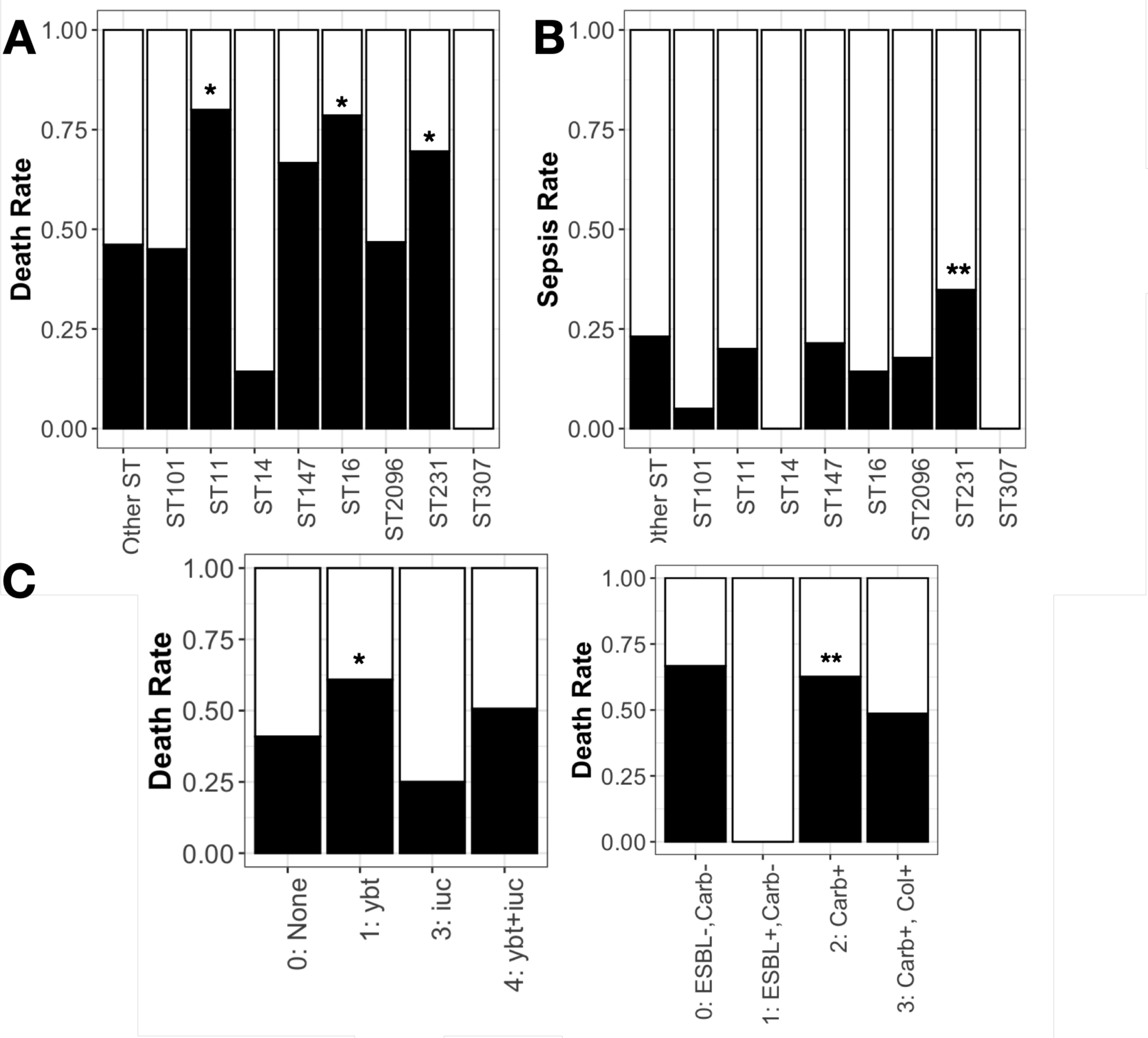
Death rate and sepsis rate for blood culture samples. A) The death rate corresponding to the ratio of patients with deceased status divided by the total number of patients with both deceased and discharged status across all STs. B) The death rate corresponding to the ratio of patients with reported sepsis in their diagnostic notes divided by the total number of patients across STs. C) The death rate corresponding to the ratio of patients with deceased status divided by the total number of patients with both deceased and discharged status across samples with different profiles of the presence of resistance/virulence genes. * and ** correspond to significance levels of 0.05 and 0.01, respectively, according to the one-sided proportion test.

**Figure S5.**
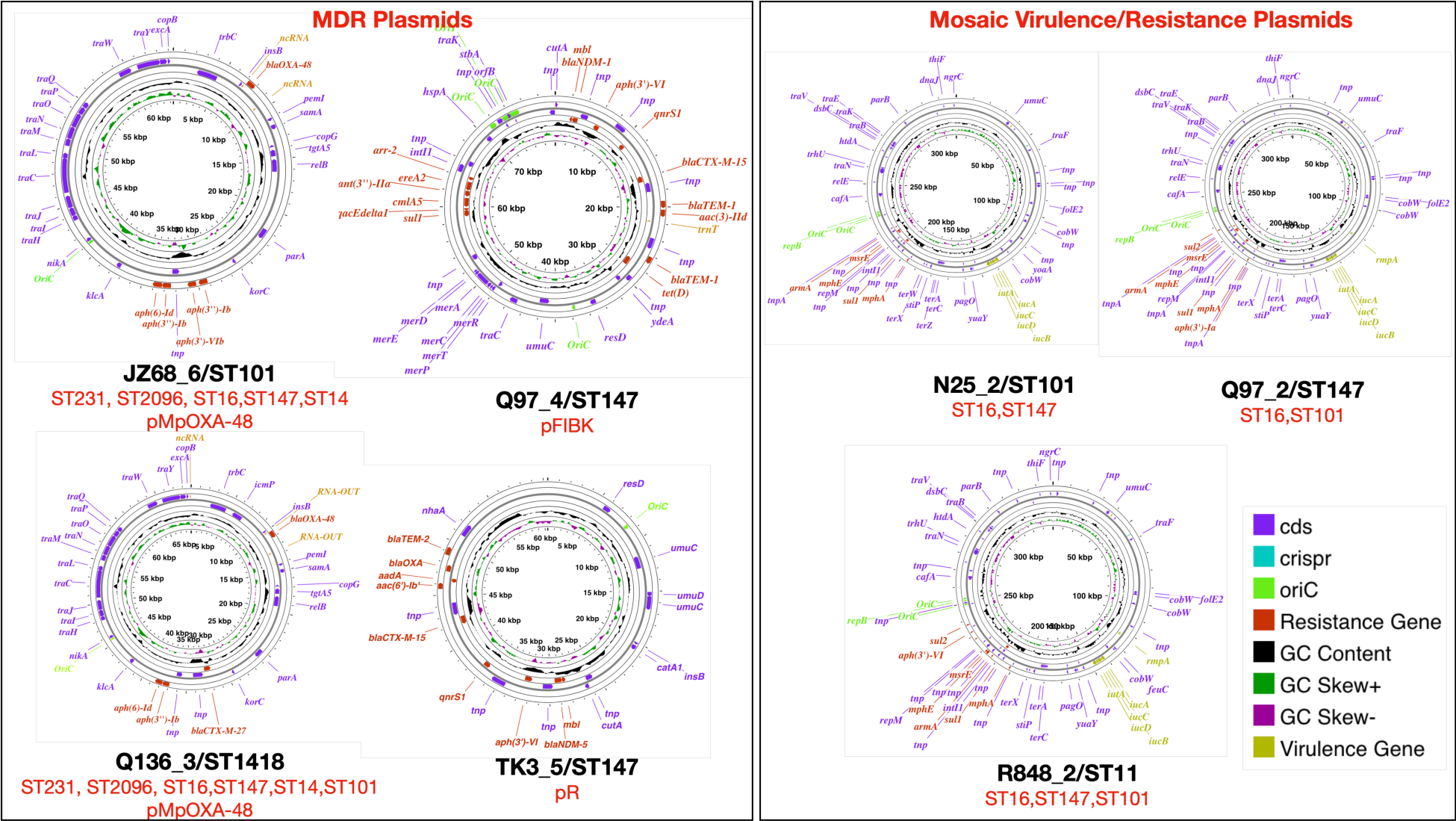
Genomic map of the MDR and virulence plasmids obtained from long-read sequencing data. The MDR plasmids depict the genetic map for plasmids carrying resistance genes and their presence in ST groups (highlighted in red text), as inferred from coverage of >90% of the reads for samples belonging to different STs. Additionally, we identified the known origin of replication for each plasmid from the PlasmidFinder database. The mosaic plasmids contain two islands for resistance and virulence genes. The complete sequences of these plasmids have been submitted to GenBank.

**Figure S6.**
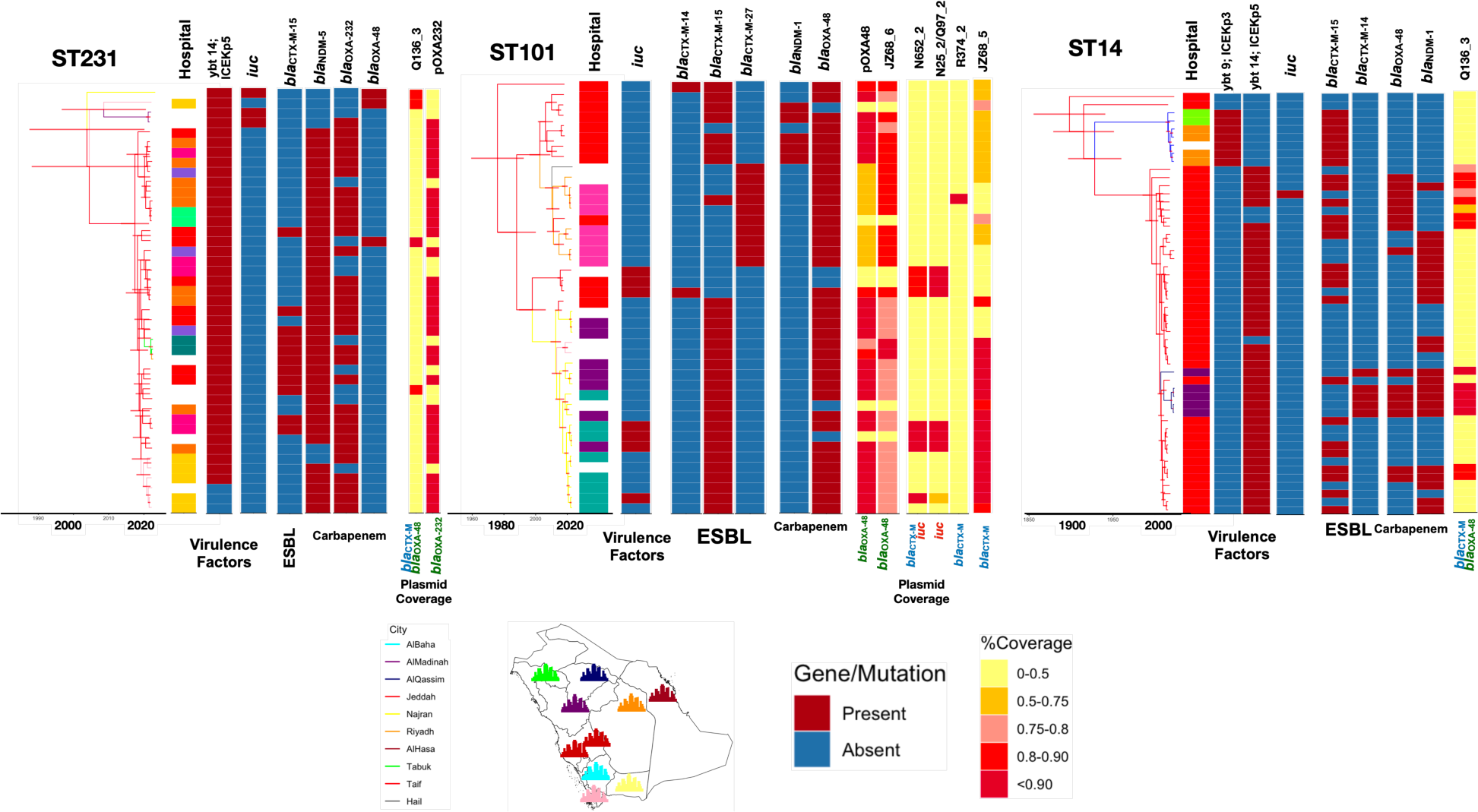
Phylogenetic trees for ST231, ST101 and ST14. The trees are Bayesian trees reconstructed by discrete state simulation. Horizontal red lines correspond to the 95% highest posterior density (HPD) interval. The colours correspond to the inferred ancestral states of the cities. The genes were identified in Kleborate. The plasmid strips show the mapping coverage of short reads against the plasmid reference genomic fragments.

